# Synaptic proteomics reveal distinct molecular signatures of cognitive change and *C9ORF72* repeat expansion in the human ALS cortex

**DOI:** 10.1101/2022.06.10.22276194

**Authors:** Zsofia I. Laszlo, Nicole Hindley, Anna Sanchez Avila, Rachel A. Kline, Samantha L. Eaton, Douglas J. Lamont, Colin Smith, Tara L. Spires-Jones, Thomas M. Wishart, Christopher M. Henstridge

**Affiliations:** Division of Cellular and Systems Medicine, School of Medicine, University of Dundee, Dundee, Scotland, UK; The Euan Macdonald Centre, Edinburgh, UK; The Roslin Institute, Royal (Dick) School of Veterinary Studies, College of Medicine and Veterinary Medicine, University of Edinburgh, Edinburgh, Scotland, UK, EH25 9RG; FingerPrints Proteomics Facility, Discovery Centre, School of Life Sciences, University of Dundee, Dundee, Scotland, UK; Academic Neuropathology, Centre for Clinical Brain Sciences, University of Edinburgh, Edinburgh, Scotland, UK; Centre for Discovery Brain Sciences and UK Dementia Research Institute at the University of Edinburgh, UK

**Keywords:** ALS, synapse, proteomics, FTD, C9ORF72, human brain

## Abstract

The two major hypotheses of Amyotrophic Lateral Sclerosis (ALS) pathogenesis (dying-forward and dying-back) have synapses at their core. Furthermore, increasing evidence suggests synaptic dysfunction is a central and possibly triggering factor in ALS. Despite this, we still know very little about the molecular profile of an ALS synapse. To address this gap, we designed a synaptic proteomics experiment to perform an unbiased assessment of the synaptic proteome in the ALS brain. We isolated synaptoneurosomes from fresh-frozen post-mortem human cortex (11 controls and 18 ALS) and stratified the ALS group based on cognitive profile (Edinburgh Cognitive and Behavioural ALS Screen (ECAS score)) and presence of a *C9ORF72* hexanucleotide repeat expansion (*C9ORF72*-RE). This allowed us to assess regional differences and the impact of phenotype and genotype on the synaptic proteome, using Tandem Mass Tagging-based proteomics. We identified over 6000 proteins in our synaptoneurosomes and using robust bioinformatics analysis we validated the strong enrichment of synapses. We found more than 30 ALS-associated proteins at the synapse, including TDP-43, FUS, SOD1 and C9ORF72. We identified almost 500 proteins with altered expression levels in ALS synapses, with region-specific changes highlighting proteins and pathways with intriguing links to neurophysiology and pathology. Stratifying the ALS cohort by cognitive status revealed almost 150 specific alterations in cognitively impaired ALS synapses, highlighting novel synaptic proteins that may underlie the synaptic vulnerability in these patients. Stratifying by *C9ORF72*-RE status revealed 330 protein alterations in the *C9ORF72*-RE+ve group, with KEGG pathway analysis highlighting strong enrichment for postsynaptic dysfunction, related to glutamatergic receptor signalling. We have validated some of these changes by western blot and at a single synapse level using array tomography imaging. In summary, we have generated the first unbiased map of the human ALS synaptic proteome, revealing novel insight into this key compartment in ALS pathophysiology and highlighting the influence of cognitive decline and *C9ORF72*-RE on synaptic composition.

## Introduction

ALS is a devastating neurodegenerative disease, caused by the loss of upper and lower motor neurons. Patients suffer progressive muscle wastage, resulting in paralysis and death, commonly within 3 years of diagnosis. There is no cure, and a greater understanding of ALS pathogenesis is required to change this bleak prognosis.

ALS is strikingly heterogenous, with 90% of cases being sporadic and the rest due to known alterations in a growing number of genes such as *SOD1, TARDBP, FUS* and *C9ORF72*. Hexanucleotide repeat expansion in *C9ORF72* (*C9ORF72*-RE) accounts for approximately 40% of familial ALS and 5% of sporadic disease, making it the most common genetic cause [1].

Growing genetic, clinical and pathological evidence suggests that ALS and Frontotemporal Dementia (FTD) lie at opposite ends of a disease spectrum. Almost one third of FTD patients have a *C9ORF72*-RE [2]. Approximately 15% of FTD patients also develop ALS and approximately 15% of ALS cases are diagnosed with co-morbid FTD [3]. In the middle of this spectrum are 30-40% of ALS patients that present with more subtle cognitive (ALS cognitive impairment (ALSci)) and/or behavioural (ALS behavioural impairment – ALSbi)) change. While these changes do not meet the criteria for FTD diagnosis, they still affect daily life and associate with quicker disease progression [4, 5]. These non-motor symptoms become much more common in the later stages of disease when up to 80% of ALS patients will present with cognitive or behavioural change [6]. Imaging studies highlight the dorsolateral prefrontal cortex (Brodmann Area 9 (BA9)) as a particularly vulnerable area in relation to cognitive decline in ALS, especially with regards to executive dysfunction which is the most common characteristic in ALSci [4, 7-11]. Furthermore, using a combination of cognitive profiling, post-mortem tissue and high-resolution imaging we recently showed that synapse loss in BA9 was associated with lower cognitive performance [12]. This was the first evidence that synapse loss was associated with cognitive change in ALS. Interestingly, recent PET studies in FTD patients revealed synapse loss in frontotemporal cortices that correlated with lower cognitive scores, thus highlighting overlapping synaptic vulnerability between ALSci and FTD [13].

Synapse loss is one of the earliest conserved pathological features across the heterogenous spectrum of ALS, occurring in the periphery at the neuromuscular junction (NMJ) and within the cortex, often before presentation of symptoms [14-16]. Recent transcriptomic and proteomic studies in human ALS brain have also highlighted changes in synaptic genes and proteins [17, 18]. However, protein alterations in small structures such as the synapse can be diluted in whole cell/tissue analyses. Furthermore, deep proteomic profiling of human brain can reveal changes in protein expression that are not highlighted by RNA expression analysis [19, 20]. This highlights the importance of using well-characterised human samples for the development of comprehensive proteomic datasets. There are no such datasets currently available for the human ALS synapse.

To address this gap, we have isolated synaptic fractions from ALS and control brain and performed high-resolution proteomic analysis to uncover the changes in the ALS synaptic proteome. We have identified almost 6000 proteins at the human synapse, showcasing the breadth of analysis possible. We provide comprehensive datasets of the synaptic proteome from two cortical brain areas, BA4 (primary motor cortex) and BA9 (dorsolateral prefrontal cortex). Furthermore, we stratified the ALS cohort by cognitive performance and *C9ORF72*-RE status to uncover the influence of these factors on the synaptic proteome. Using a robust bioinformatics approach, we have uncovered specific pathways and proteins altered in the ALS synaptic proteome compared to controls. Importantly, we discovered the presence of a *C9ORF72*-RE has a distinct effect on the synaptic proteome.

This study not only provides valuable datasets for the field, but also strengthens the hypothesis for synaptic dysfunction as an important feature of ALS and highlights proteins and pathways that could be exploited for future therapeutic development.

## Materials and Methods

### Donor characteristics and brain collection

All donors were assessed using the revised El Escorial criteria for diagnosing ALS [21]. Patients were recruited through the Scottish Motor Neurone Disease Register and data collected in the CARE-MND database (Clinical Audit Research and Evaluation). Ethical approval for this register was obtained from Scotland A Research Ethics Committee 10/MRE00/78 and 15/SS/0216. All clinical data were subsequently extracted from the CARE-MND database. Use of patient samples for genetic profiling has been approved by the Chief Scientist Office Scotland; MREC/98/0/56 1989–2010, 10/MRE00/77 2011–2013, 13/ES/0126 2013–2015, 15/ES/0094 2015-present.

The genetic and cognitive status of ALS patients was obtained via the CARE-MND database. Cognition was assessed using the Edinburgh Cognitive and Behavioural ALS Screen (ECAS, [22]) and genetic profile confirmed as previously described [23]. Most of the cases were cognitively and genetically profiled for our previous study [12]. Following completion of the proteomics data it became apparent that one ALS case was incorrectly classified as ALSci and in fact had a normal ECAS score (red symbol in Supplementary Figure 1D). We expect this to have minimal effect as the influence of this one sample will be diluted by the other 8 within the ALSci experimental groups. Whether we physically pool individual samples or generate a mean value from individual samples after western blot, the result is remarkably similar, suggesting the influence of a single sample is minimal on the overall group result (Supplementary Figure 3).

Use of human tissue for post-mortem studies has been reviewed and approved by the Edinburgh Brain Bank ethics committee and the ACCORD medical research ethics committee, AMREC (ACCORD is the Academic and Clinical Central Office for Research and Development, a joint office of the University of Edinburgh and NHS Lothian, approval number 15-HV-016). The Edinburgh Brain Bank has research ethics committee (REC) approval (21/ES/0087).

Fresh frozen and formalin-fixed brain tissue was obtained from the primary motor cortex (BA4) and the dorsolateral prefrontal cortex (BA9) of age/gender matched clinically diagnosed ALS patients (n=11) and control (no neurological condition) individuals (n=18). Summary demographics can be found in Table 1.

**Table 1:** Summary demographic data of donors. Post-mortem interval, age at death and ECAS score are presented as mean ± S.D. ECAS score was significantly lower in ALSci compared to ALSnoci (* 2-tailed t-test; p=0.0005). No differences were found between any other group comparisons.

| Experimental Group | n | Post-mortem Interval (hrs) | Age at death (years) | ECAS Score (Max 136) | Gender (% Male) |
| --- | --- | --- | --- | --- | --- |
| Control | 11 | 74 ± 24 | 66 ± 18 |  | 64 |
| ALS | 18 | 81 ± 29 | 64 ± 10 |  | 56 |
| ALSnoci | 9 | 95 ± 29 | 66 ± 9 | 117 ± 5 | 44 |
| ALSci | 9 | 67 ± 24 | 62 ± 11 | 99 ± 11* | 67 |
| ALS c9-ve | 12 | 88 ± 29 | 58 ± 9 | 108 ± 15 | 58 |
| ALS c9+ve | 6 | 66 ± 27 | 67 ± 9 | 108 ± 8 | 50 |

### Preparation of synaptoneurosomes

Total homogenate (TH) and synaptoneurosome (SNS) preparation was performed as described previously [24, 25]. Briefly, tissue from BA4 and BA9 from each case was homogenised in homogenisation buffer (25mM HEPES (pH 7.5), 120mM NaCl, 5mM KCl, 1mM MgCl_2,_ 2mM CaCl_2_, protease inhibitors (Roche #11836153001) and phosphatase inhibitors (Thermo Scientific #A32959)) using glass homogenisers, on ice. The homogenised tissue was then filtered through an 80µm nylon filter, and a TH aliquot was stored on dry ice. The remainder of the crude homogenate was filtered further through a 5µm filter (Millipore, SLSV025LS) and centrifuged for 5 minutes at 1000g. Pellets were washed in homogenisation buffer and supernatant was disposed. The final pellet was weighed and resuspended in 100mM Tris-HCL Buffer (pH 7.6) 4% SDS containing 1% protease inhibitor cocktail (Roche #11836153001) in 1:5 dilution based on pellet weight. Samples were further homogenised by hand and centrifuged at 17,000g at 4°C for 20 minutes, and the supernatant collected. Concentration of the samples were determined using a Micro BCA Protein Assay kit (Thermo Scientific #23235). 25µg of protein from each individual donor was pooled based on the stratification of the subjects (Supplementary Figure 1A), to generate 10 experimental pools. This resulted in one representative pooled sample for each group, along with original samples from individuals, and all samples were stored at -80°C until further experiments.

### Western Blot

10µg of sample was prepared with 4X Laemilli sample buffer (Bio-Rad, #1610747), beta-mercaptoethanol and denatured at 95°C for 5 minutes. Samples were loaded into 4-20% Tris-Glycine 1.0mm polyacrylamide precast gels (Thermo Fisher, #WXP42020BOX) along with 5µl protein ladder (Li-Cor #928-70000). After electrophoresis, proteins were transferred onto nitrocellulose membrane using precast transfer stacks (Invitrogen, #IB23001) with the iBlot™ 2 Gel Transfer Device (Invitrogen, #IB21001). Membrane was then stained for total protein using REVERT™ 700 Total Protein stain (Li-Cor, REVERT™ 700 Total Protein Stain Kits, #926-11010) and imaged with the Li-Cor Odyssey system. After imaging, the membrane was destained based on manufacturer’s instructions. Membranes were block with 5%milk/TBST or 5% BSA/TBST solution for an hour. Primary antibodies were diluted in the suitable block solution and membranes were incubated overnight at 4°C. Next day, after extensive washes secondary antibody was diluted in block and incubated for an hour at room temperature, then imaged with the Li-Cor Odyssey system. The details of primary and secondary antibodies are summarized in Supplementary Table 1. Total protein stains shown in Supplementary Figure 1D were generated by running 3ug total protein in 4-20% Tris-Glycine 1.0mm polyacrylamide precast gels (as above) and performing an Instant Blue total protein stain, per manufacturer instructions (Expedeon).

### Tandem Mass Tagged (TMT) Liquid Chromatography Mass Spectrometry (LC-MS/MS)

#### S-Trap processing of samples

100µg of total protein per experimental group was processed using S-trap mini protocol (Protifi) as recommended by the manufacturer. Samples were applied on the S-trap mini spin column and trapped proteins were washed 5 times with S-TRAP binding buffer. Samples were digested with trypsin (1:50) overnight at 37°C in 150µl of TEAB at a final concentration of 100mM. Peptides from S-Trap mini spin column were eluted by centrifugation at 4000g for 30 seconds in 160 µl of 50 mM TEAB, then 160 µl of 2% aqueous formic acid and finally 160 µl of 50% acetonitrile/0.2% formic acid. Resulting tryptic peptides were pooled, dried and quantified using Pierce Quantitative fluorometric Peptide Assay (Thermo Scientific).

#### 10-plex TMT Labelling and high pH reverse phase fractionation

Samples were labelled with TMT tags using Pierce High pH Reversed-Phase Peptide Fractionation kit (Thermo Scientific, #84868) following manufacturer’s protocol. Desalted tryptic peptides (37µg each sample) were dissolved in 100µl of 100mM TEAB. The 10 TMT labels were added to different samples after being dissolved in 41µl of anhydrous acetonitrile. Mixtures were incubated for 1 hour at room temperature, 8µl of 5% hydroxylamine was added per sample to stop the labelling reaction. Samples were mixed, desalted and dried in speed-vac at 30°C following labelling with TMT. 200µl of ammonium formate (NH₄HCO₂) (10mM, pH 9.5) was used to re-dissolve the samples and peptides were fractionated using High pH RP Chromatography. A C18 Column from Waters (XBridge peptide BEH, 130Å, 3.5 µm 2.1 × 150 mm, Waters, Ireland) with a guard column (XBridge, C18, 3.5 µm, 2.1×10mm, Waters) were used on an Ultimate 3000 HPLC (Thermo Scientific). Buffer A: 10mM ammonium formate in milliQ water pH 9.5 and Buffer B: 10mM ammonium formate, pH 9.5 in 90% acetonitrile were used for fractionation. Fractions were collected at 1 minute intervals using a WPS-3000FC auto-sampler (Thermo Scientific). Column and guard were equilibrated with 2% Buffer B for 20 minutes at a flow rate of 0.2ml/min. Separation gradient of column was started 1 minute after 190µl of TMT labelled peptides were injected into the column. Elution of peptides was done with a column gradient of 2% Buffer B to 10% Buffer B in 6 minutes, and then from 10% Buffer B to 47% Buffer B in 53 minutes. Column was washed for 15 minutes in 100% Buffer B and equilibrated at 2% Buffer B for 20 minutes. Fraction collection resulted in a total of 80 fractions, 200µl each, as the fraction collection was stopped after 80 minutes. The total number of fractions concatenated was set to 20 and the content of the fractions was dried and suspended in 50µl of 1% formic acid prior to analysis with LC-MS.

#### LC-MS analysis

Mass spectrometry analysis was carried out at the ‘FingerPrints’ Proteomics Facility, School of Life Sciences, University of Dundee.

Analysis of peptides was performed on a Q-Exactive-HF (Thermo Scientific) mass spectrometer coupled with a Dionex Ultimate 3000 RSLC Nano (Thermo Scientific). LC buffers were the following: buffer A (0.1% formic acid in Milli-Q water (v/v)) and buffer B (80% acetonitrile and 0.1% formic acid in Milli-Q water (v/v). Aliquots of 5-7.5 μL of each sample were loaded at 10 μL/min onto a trap column (100 μm × 2 cm, PepMap nanoViper C18 column, 5 μm, 100 Å, Thermo Scientific) equilibrated in 0.1% formic acid. The trap column was washed for 5 min at the same flow rate with 0.1% formic acid and then switched in-line with a Thermo Scientific, resolving C18 column (75 μm × 50 cm, PepMap RSLC C18 column, 2 μm, 100 Å). The peptides were eluted from the column at a constant flow rate of 300 nl/min starting from 5% buffer B to 5% buffer B in 8 min, then from 5% buffer B to 35% buffer B in 125 min, and then to 98% buffer B within 2 min. The column was then washed with 98% buffer B for 20 min and re-equilibrated in 5% buffer B for 17 min. The column was kept all the time at a constant temperature of 50°C.

Q-Exactive HF was operated in data dependent positive ionisation mode. The source voltage was set to 2.4 kV and the capillary temperature was 250°C. A scan cycle comprised MS1 scan (m/z range from 335-1600, with a maximum ion injection time of 50 ms, a resolution of 120000 and automatic gain control (AGC) value of 3×10^6^) followed by 15 sequential dependant MS2 scans (resolution 60000) of the most intense ions fulfilling predefined selection criteria (AGC 1×10^5^, maximum ion injection time 200 ms, isolation window of 0.7 m/z, fixed first mass of 100 m/z, NCE/Stepped nce32, spectrum data type: centroid, AGC target of 1e5, exclusion of unassigned, singly and >6 charged precursors, peptide match preferred, exclude isotopes on, dynamic exclusion time of 45 s). Mass accuracy is checked before the start of samples.

#### Peptide Quantification

The raw mass spectrometric data files obtained for each experiment were collated into a single quantitated data set using MaxQuant (version 1.6.2.10) [26] and searched against the SwissProt subset of the *H. sapiens* Uniprot database (May 2019 release) using the Andromeda search engine software [27]. Enzyme specificity was set to that of trypsin, allowing for cleavage of N-terminal to proline residues and between aspartic acid and proline residues. Other parameters used were: (i) variable modifications, methionine oxidation, deamidation (N,Q) protein N-acetylation, gln - pyro-glu, Phospho(STY); (ii) fixed modifications, cysteine carbamidomethylation; (iii) TMT 10-plex labels; (iv) MS/MS tolerance: FTMS-10ppm, ITMS-0.06 Da; (vi) maximum peptide length, 6; (vii) maximum missed cleavages, 2; (vii) maximum of labelled amino acids, 3; and (viii) false discovery rate, 1%. The correction factors for the TMT labelling were also applied. MS RAW files were uploaded to the PRIDE proteomics repository (https://www.ebi.ac.uk/pride/) with the following dataset ID **(to be updated once published)**.

### Bioinformatic analyses

MaxQuant output files were filtered by number of unique peptides per protein and by missing values. Protein identifications that were assigned with ≥2 unique peptides were retained. In addition, those proteins which were not detected in all samples, were excluded. After these robust clean-up steps, there were 5,348 analysis-ready proteins. ALS data was normalised to control and then 1/median normalized to correct for any minor loading variables, generating ratiometric values for each protein for each experimental comparison. We chose a threshold vale of 20% as a relevant change in protein expression (≥1.2 or ≤0.8 ratio change). This value was chosen as post-hoc validation of protein change by modern western blotting technology is sensitive enough to validate these changes [25, 28]. This analysis-ready dataset can be found in Appendix 2.

#### Enrichment analysis

All Swiss-Prot ID were inputted into Database for Annotation, Visualisation and Integrated Discovery (DAVID) software to identify enriched biological themes within the dataset [29, 30]. This database generates a gene-to-gene similarity matrix using a clustering algorithm to classify highly related genes into functionally related groups. As DAVID can only analyse a maximum of 3000 IDs at once, we halved our dataset using a random number generator tool in Excel. Each protein ID was assigned a random number which was then organised by descending order. Group1 and Group2 contained 2674 gene symbols (Supplementary Figure 2D). Clusters were identified using the Functional Annotation Clustering tool. For more information on the statistics and enrichment algorithms, see the original paper and recent update [29, 30].

To identify the enriched Gene Ontology terms (GO) and KEGG pathways we used g:Profiler and ShinyGo v0.75. Swiss-Prot IDs were submitted into the online platforms with the specification of *Homo Sapien* datasets. Then, enriched terms were downloaded and plotted.

#### Data visualization

For visualization of enrichment data we used R version 4.0.5, ggplot2 package. For calculating and plotting Venn diagrams, we used the free online software of the Van de Peer Lab: https://bioinformatics.psb.ugent.be/webtools/Venn/. For visualizing heatmaps, first, all protein ratiometric values were log2 transformed, and the Z-score was counted using the following formula:

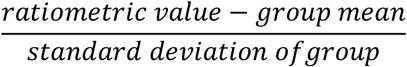

Heatmaps were plotted using the heatmap.2 module from the gplots package of R software, using hierarchical clustering with Euclidean distance.

Workflow and schematic figures were prepared using BioRender, www.biorender.com

### Array Tomography

Tissue was processed for array tomography as previously described [31]. Briefly, fresh post-mortem tissue from BA9 was dissected and fixed in 4% paraformaldehyde in PBS for 2-3h. Samples were then dehydrated through ascending ethanol washes (50%, 70%, 90% and 100%, respectively) and incubated in LR White resin (Electron Microscopy Sciences) overnight. Blocks were then placed individually into capsules containing LR White and polymerized overnight at 60⁰C. Tissue blocks were cut into serial sections of 70nm thickness using an ultracut microtome (Leica UC7) with a Histo Jumbo Diamond knife (Diatome, Hatfield, PA). Tissue ribbons were collected on gelatin-coated glass coverslips and permeabilised using a 50mM glycine solution and blocked for 30min (0.1% fish skin gelatin and 0.05% Tween20 in TBS). Afterwards, samples were immunostained with primary antibodies against synaptophysin (1:50, ab8049, Abcam) and synaptopodin (1:50, 21064-1-AP, Proteintech) overnight. No primary, negative control was included to rule out non-specific binding of the secondary antibodies. The following day, the staining was developed with fluorescently-labelled secondary antibodies (1:50 donkey anti-mouse Alexa fluor 488 – ab150105, Abcam; 1:50 donkey anti-rabbit Alexa fluor 594 – ab150076, Abcam, respectively) for 30 min and DAPI (1:1000, ab228549, Abcam) for 5 min. Samples were then mounted onto slides with Immumount mounting media. Images were obtained at the same position on each section along the ribbon using a DeltaVision Elite widefield fluorescence microscope (Image solutions) equipped with a CoolSnap digital camera and softWoRx software. High resolution images were obtained with a 63x 1.4NA Plan Apochromat objective. At least two image stacks were captured per region for each case. Stacks were aligned using the ImageJ Multistack Reg plugin [32]. All aligned image stacks were then thresholded, segmented and puncta densities measured using a custom MATLAB script. Colocalisation analysis of the segmented images was performed using an in-house Python script. All custom software can be downloaded from GitHub at https://github.com/arraytomographyusers/Array_tomography_analysis_tool and https://github.com/lewiswilkins/Array-Tomography-Tool.

## Results

To ensure the production of a high-quality proteomics dataset that allows biologically meaningful comparisons to be made, we designed an experiment comprising 29 brain donors (11 healthy controls and 18 ALS cases). Samples were age and gender matched and summary demographic information can be found in Table 1.

Our experimental design incorporated disease, brain region, *C9ORF72*-RE status and cognitive profile, as summarised in Supplementary Figure 1A. To prevent potential confounding influences of subtle differences in brain handling, sample preparation and features unique to an individual, we pooled samples into 10 pre-determined groups (Supplementary Figure 1A). Pooling produces a representative sample for each condition, consisting of numerous individual cases which can be analysed for inter-individual variability during later validation work [25, 28]. We have retained all individual samples for future validatory experiments. Furthermore, it allows the direct comparison of multiple conditions at the same time in the same mass-spectrometer, eliminating the natural variability accompanying multiple experimental runs.

The quality of the synaptically-enriched preparations (Supplementary Figure 1C-E) was assessed using well-characterised molecular approaches as described previously [24, 25, 28]. We first confirmed that the nuclear protein lamin was not detected in synaptic preparations and the synaptic protein synaptophysin was enriched (Supplementary Figure 1C). Furthermore, in addition to the 10 experimental pools, all constituent individual samples were run as total protein gels and consistent banding and expression between samples was observed (Supplementary Figure 1D,E). We have shown in previous work using transmission electron microscopy that intact synapses with clear pre- and postsynaptic compartments are retained in these synaptoneurosome preparations [24, 25].

Following the validation of synaptoneurosome quality, we initiated a 10-plex TMT LC-MS/MS proteomics workflow (Figure 1A, Supplementary Figure 1B). In total, 6059 proteins were putatively detected. We filtered these protein IDs based on the stringent identification criteria of 2 or more unique peptides and removed any proteins not detected in one or more groups, yielding 5348 analysis-ready proteins (Supplementary Figure 2A). Therefore, all proteins analysed throughout were identified by ≥2 unique peptides and identifiable within all 10 samples, allowing robust comparisons of protein expression to be made between groups. To confirm that our dataset profiled an enriched synaptic proteome, we performed several alignment and enrichment analyses. Firstly, we aligned our filtered dataset with two recent human brain and synapse proteome studies [33, 34]. This revealed >99% of our dataset matched recently published data (Figure 1B). We also aligned our data with three synapse proteome databases [35-37], revealing >90% of our data overlapped (Supplementary Figure 2B). KEGG pathway analysis showed strong enrichment of common neuronal and synaptic pathways (Figure 1C) and ShinyGO online analyses [38] revealed strong enrichment of synaptic terms in cellular compartment, molecular function, and biological processes (Figure 1D). Utilising two further enrichment tools (g:Profiler [39] and DAVID functional annotation [29]), we continually reported clear enrichment of synaptic terms (Supplementary Figure 2C,D).

**Figure 1:**
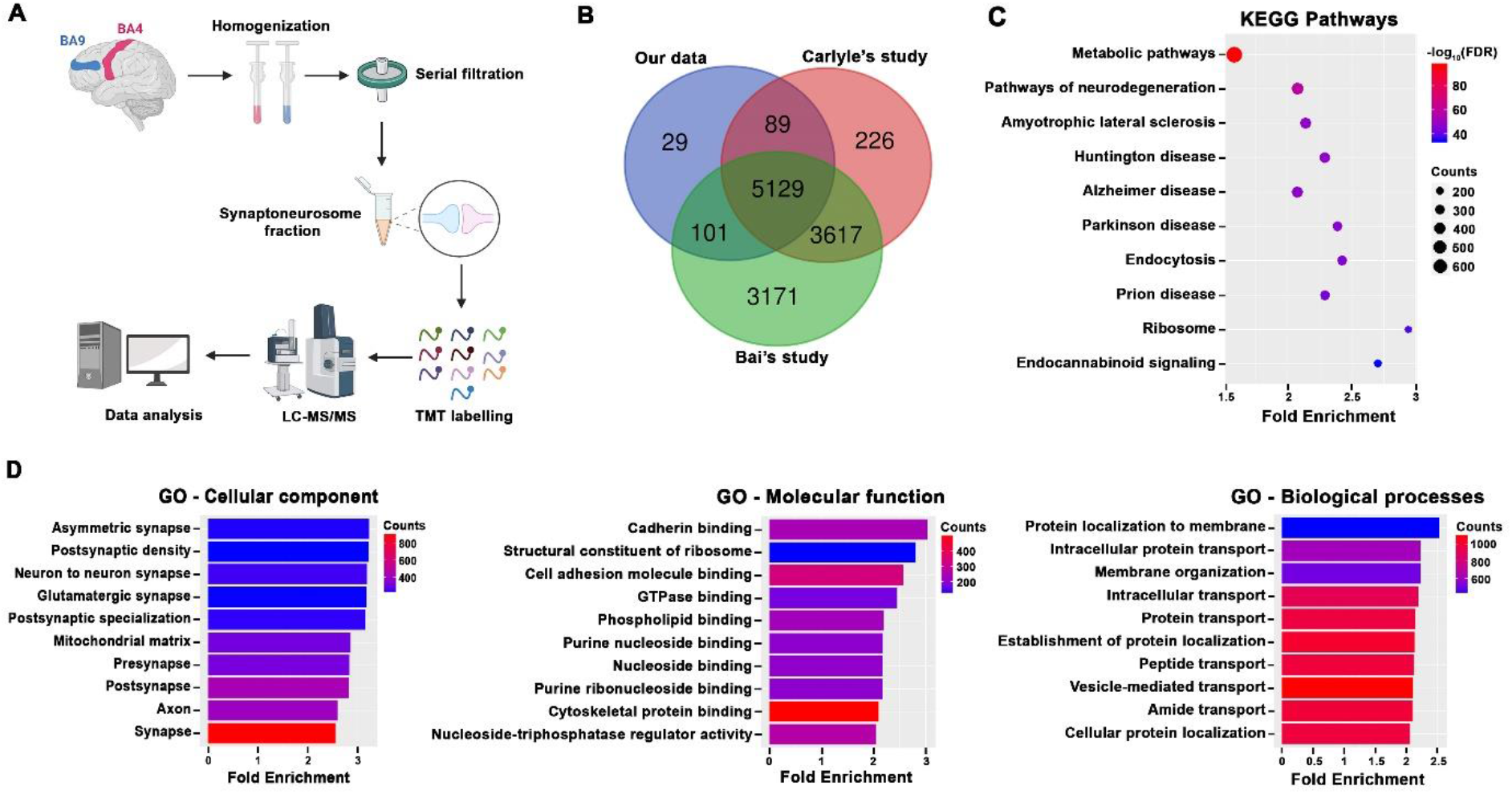
Enrichment analysis of our synaptic fractions. **A**. Schematic diagram shows experimental workflow. Synaptically-enriched fractions were collected using BA9 and BA4 brain areas from both control and ALS patients. Synaptic protein fractions were Tandem Mass Tag (TMT) labelled and analysed by liquid chromatography, mass spectrometry-based quantification and identification (LC-MS/MS). Data was analysed using MaxQuant quantitative proteomics software. **B**. The identified 5348 unique proteins demonstrate a 99% overlap with previously published human proteomics data. **C-D**. Bioinformatics analysis using ShinyGo enrichment analyser with protein IDs show the top 30 KEGG pathways (**C**) and plotted based on fold enrichment and gene counts/pathway (dot size), colorized by -log_10_(FDR). Colorized bar graphs by gene counts show (**D**) the top 15 enriched biological terms in Gene Ontology (GO) clusters.

Taken together, this comprehensive analysis revealed strong enrichment of synaptic components in our samples. To ensure changes identified by mass-spectrometry truly reflect the protein changes within the samples, we assessed the change in expression of a selection of proteins by western blot. Ratiometric change in protein expression was quantified (by dividing ALS protein intensity by control intensity) and plotted against the proteomics ratiometric data (by dividing ALS protein intensity by control intensity and correcting by 1/median), revealing a robust correlation (R^2^= 0.849, p=0.0032; Supplementary Figure 3A,B). To assess inter-individual variability within our group samples, we randomly selected 6 ALS samples and 6 controls, then blotted for two proteins (Supplementary Figure 3C). While inter-individual variability was high, the mean change in expression closely matched the proteomics data, showing increases and decreases where expected (Supplementary Figure 3C).

Confident that we have a synaptically-enriched dataset and a validated proteomics approach, we first looked for any ALS-associated proteins within our data. Using four recently published studies [1, 40-42] we collated 58 ALS-associated genes and found the protein product of 37 of them in our dataset (Supplementary Figure 4A). Proteins were described to have altered abundance if their expression changed by more than 20%. Importantly, we see expression changes in some of these proteins including FUS, SOD1, SQSTM1 and MATR3. As observed in previous work [43] we see a remarkably similar 20% decrease in expression of C9ORF72 in the *C9ORF72*-RE+ve group (Supplementary Figure 4B). The most significant change is an upregulation of COG3 (part of the Conserved Oligomeric Golgi complex) in ALS BA4 and BA9. COG3 is a recently identified ALS-associated gene [40] and its role is yet to be elucidated in ALS. COG proteins are essential for regulating Golgi processes and influence protein trafficking and glycosylation in neurons, with mutations linked to several neurological disorders [44]. Golgi fragmentation occurs presymptomatically in models of ALS and is observed in both familial and sporadic human patients (reviewed in [45]). Interestingly, several members of the COG complex were found to be altered in our dataset, with some increased across all groups (Supplementary Figure 4C,D) suggesting Golgi satellites near synapses are disrupted in ALS.

Following this targeted analysis of ALS-associated proteins, we next took a broader look at synaptic protein change between control and ALS samples from both BA4 and BA9. We identified almost 500 proteins in either BA4 or BA9 with altered expression in ALS compared to controls (Supplementary Figure 5A).

When stratified by region, which revealed region-specific changes in protein abundance (Figure 2A), more than 300 proteins changed in ALS BA4 (236 UP, 82 DOWN) and almost 250 changed in ALS BA9 (168 UP, 82 DOWN) when compared to controls (Supplementary Figure 5A). KEGG pathway analyses using just the up- and downregulated proteins revealed enrichment for inflammatory pathways such as complement and coagulation pathways in both brain areas (Figure 2B,C). Interestingly, BA4 revealed more diverse changes at the pathway level, highlighting the importance of GABAergic signalling and RNA splicing in ALS (Figure 2B). Following further analysis using the synaptic database by Sorokina and colleagues [35] we discovered that most of the altered proteins were postsynaptic in both brain areas (Figure 2D). Of the >300 altered proteins in BA4, 221 were specific for that region and KEGG analysis revealed strong enrichment of lipid metabolism pathways (Figure 2E-G). Given the strong postsynaptic enrichment of protein alterations in Figure 2D, we submitted the BA4-specific proteins into the experimentally validated SynGO database, which revealed the subsynaptic localization of each protein (Figure 2J). Despite a small percentage of presynaptic change (Figure 2D), SynGO highlighted disruption in presynaptic vesicular trafficking (Figure 2J). Postsynaptic glutamate receptor upregulation and cytoskeletal changes dominated, which might reflect postsynaptic adaptation to the presynaptic malfunction of vesicular release.

**Figure 2.**
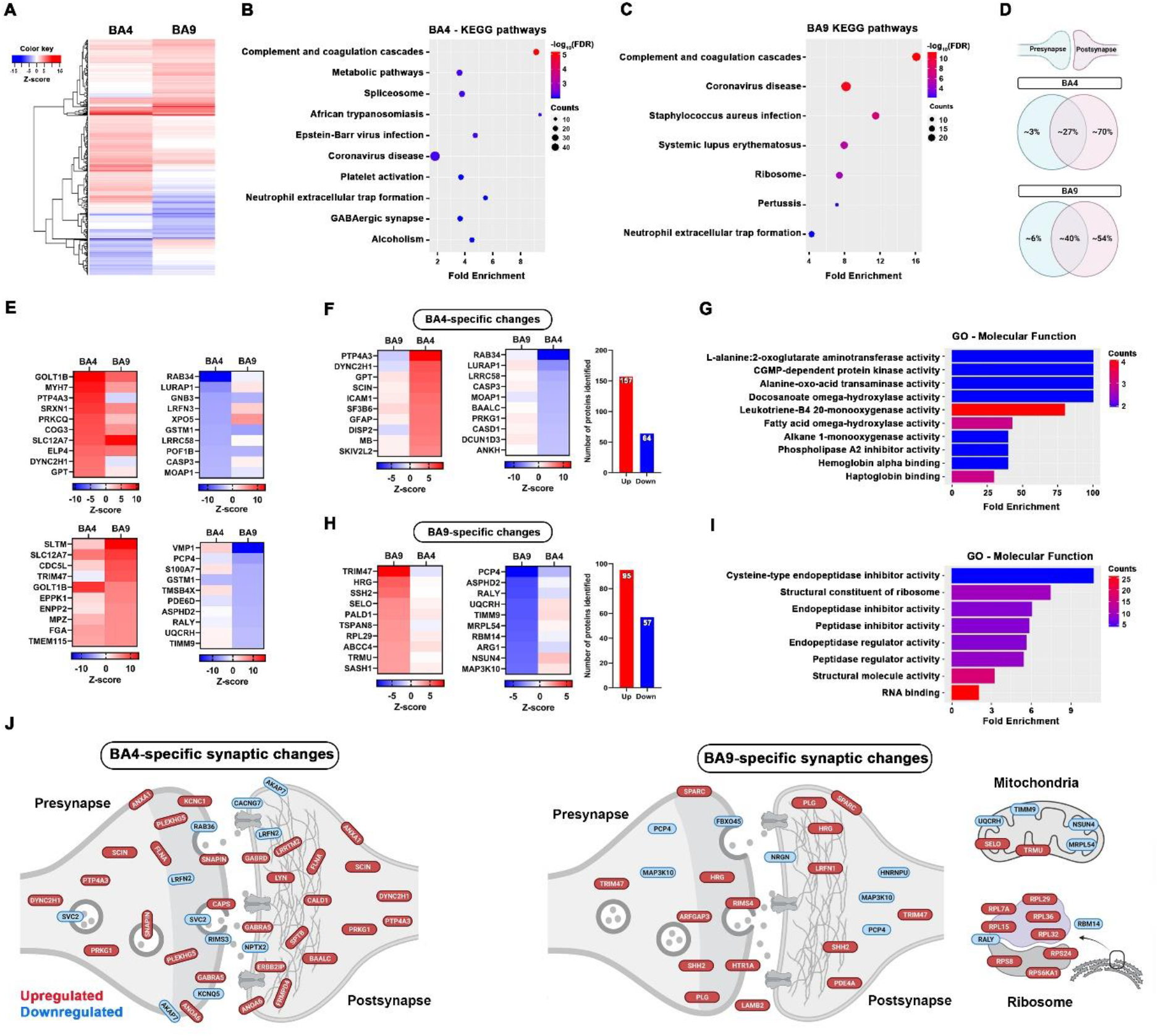
Regional differences in the ALS synaptic proteome. **A**. Heatmap shows the region-specific protein changes. KEGG pathways show the most enriched terms in both BA4 (**B**) and BA9 (**C**) using only the ≥20% changed protein IDs. Graphs were plotted based on fold enrichment and gene counts/pathway (dot size) and coloured based on -log10(FDR) **D**. Subsynaptic compartmentalization of the changed proteins utilising the Sorokina database shows mostly postsynaptic enrichment in both brain areas. **E**. Top 10 up- and downregulated proteins in both brain areas. **F**,**H**. Top up- and downregulated proteins specifically changed in highlighted brain area. **G**,**I**. Enrichment analysis shows the top GO - molecular function terms in both brain areas. **J**. Subsynaptic localization of the brain area-specific, up- and downregulated proteins, discovered by the SynGO database. **D** and **J** were created with BioRender.com.

In BA9, 152 proteins were specifically altered, which were mostly enriched in peptide metabolism and synaptic translation pathways (Figure 2H, I). As can be seen from Figure 2J, we also found a remarkable downregulation of key mitochondrial proteins which may reflect mitochondrial stress and energy disbalance at the synapse (Figure 2J). Several ribosomal proteins were specifically upregulated at the BA9 synapse, possibly highlighting a dysfunction in synaptic RNA processing as an early marker of ALS synaptic stress.

ALS patients with cognitive impairment (ALSci) have a worse disease prognosis [46] and the underlying pathophysiology for this is unknown. We previously discovered synaptic loss associated with cognitive decline in BA9 in ALSci [12], however, nothing is known about the molecular changes at the synapse. To address this, we analysed the synaptic proteome in BA4 and BA9 in ALSci samples (n=9) versus ALS cases with no cognitive impairment (ALSnoci; n=9). We discovered altered expression of more than 400 proteins in ALS BA9 compared to control, when samples were stratified by cognitive status (Supplementary Figure 5C, Figure 3A). Enriched KEGG pathways within the altered proteins include numerous pathways involved in molecular transport and protein localisation and complement pathways (Figure 3B). More than 260 proteins changed in the ALSci group compared to controls and 143 of these were specific for the ALSci group, 100 proteins were upregulated and 43 downregulated (Fig 3C-E). Gene ontology analysis of these 143 proteins revealed enrichment of inflammatory signalling pathways and RNA biology (Figure 3E). In addition, subsynaptic compartmentalization analysis revealed that important synapse-spanning adhesion (CDH7) and postsynaptic scaffolding proteins (FRMPD4) are downregulated, meanwhile several ribosomal proteins are upregulated, possibly in an attempt to bolster deteriorating synaptic protein homeostasis (Figure 3F).

**Figure 3.**
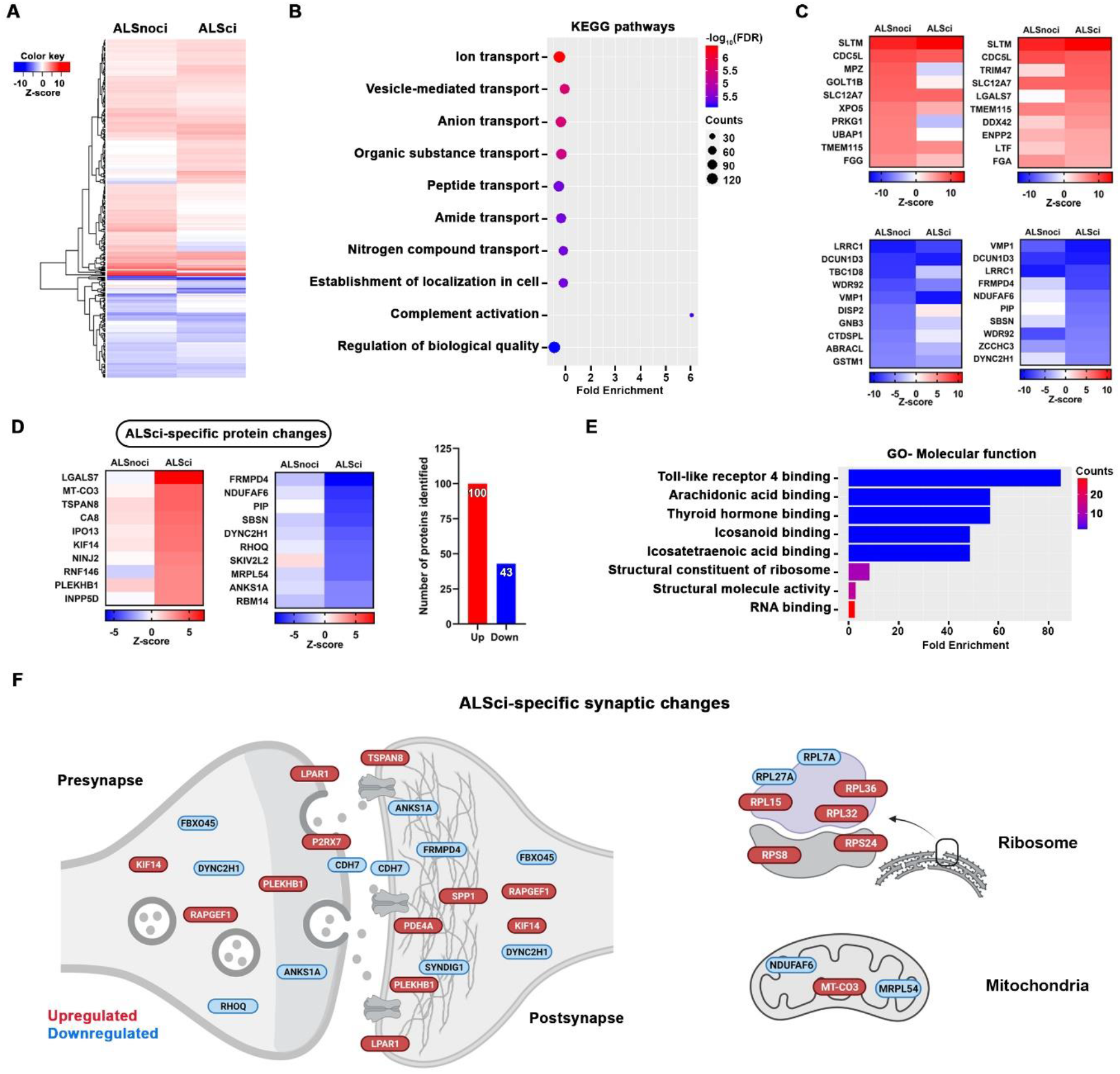
Cognitive impairment is associated with specific pathway changes in BA9. **A**. Heatmap shows the protein expression differences between cognitively impaired (ALSci) and non-impaired (ALSnoci) groups. **B**. KEGG pathway analysis visualises the most enriched pathways in these groups. Graph was plotted based on fold enrichment and gene counts/pathway (dot size), coloured based on -log10(FDR). **C**. Heatmaps show the Top 10 up- and downregulated proteins in each group. **D**. Top 10 ALSci-specific protein changes with the number of the up and down-regulated proteins. **E**. Gene ontology analysis show the most enriched molecular function terms using ALSci-specific proteins. **F**. Subsynaptic localization of the ALSci-specific protein changes, discovered by the SynGO database. Schematic figure was created by using BioRender.com.

In BA4 we found a more severe change with over 600 proteins altered across ALSci and ALSnoci groups compared to controls (Supplementary Figure 5B, Figure 4A). KEGG pathway analysis of these proteins revealed enrichment of pathways involved in complement activation, metabolic pathways and the regulation of actin cytoskeleton (Figure 4B). When stratified by cognitive status we found that ALSci BA4 synapses expressed 271 upregulated and 114 downregulated proteins (Supplementary Figure 5B, Figure 4C). ALSci-specific synaptic changes in BA4 consisted of 162 upregulated and 68 downregulated proteins (Figure 4D). Gene ontology of these 230 ALSci-specific proteins in BA4, revealed significant enrichment of RAGE receptor biology, cytoskeleton pathways and RNA biology (Figure 4E). Interestingly, subsynaptic analysis identified proteins involved in synaptic vesicle docking and turnover were downregulated, along with several key proteins involved in synaptic adhesion (Figure 4F).

**Figure 4.**
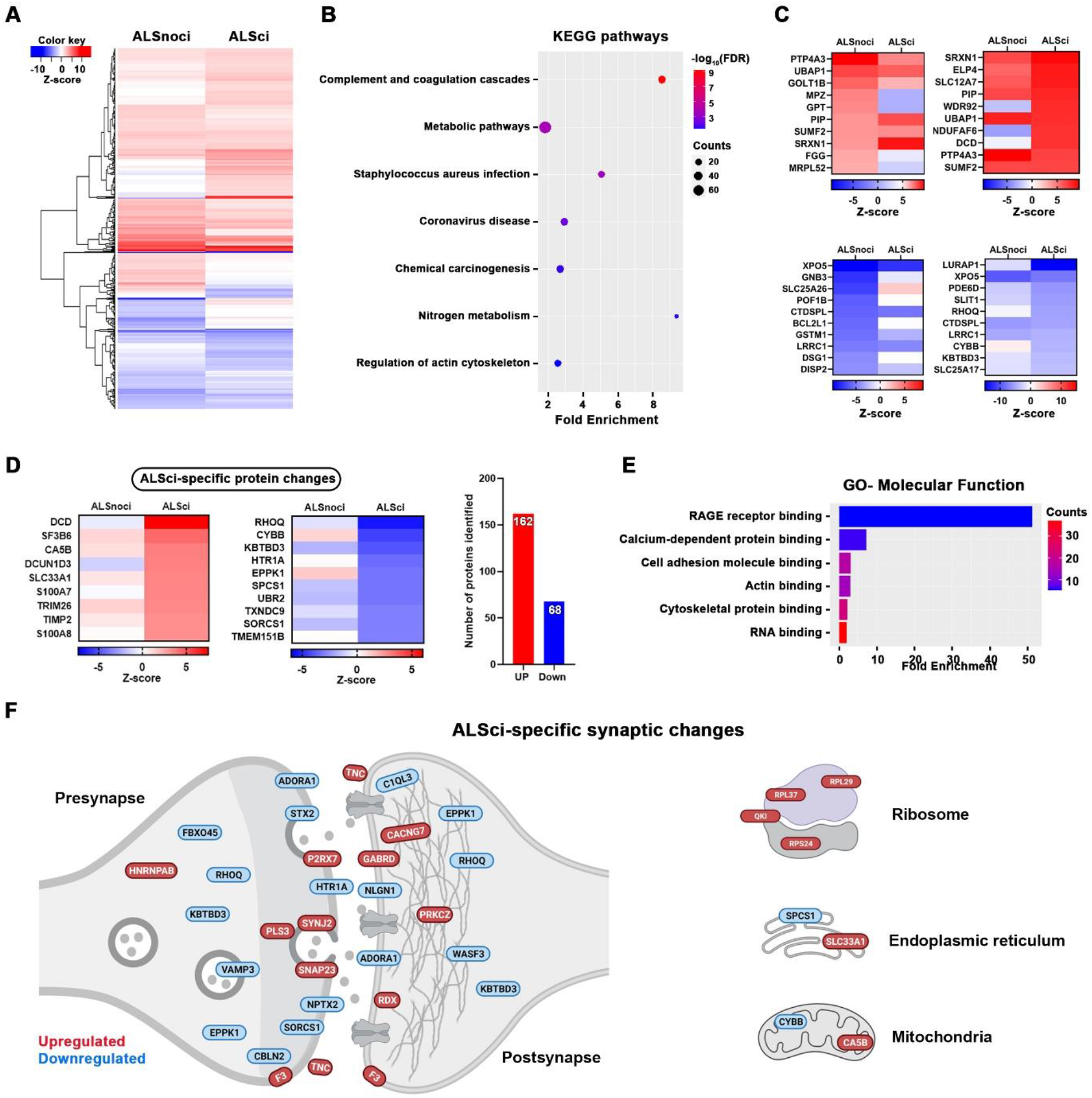
Cognitive impairment is associated with specific pathway changes in BA4. **A**. Heatmap shows the protein expression differences between non cognitively impaired (ALSnoci) and impaired (ALSci) groups. **B**. Further enrichment analysis shows the most enriched KEGG pathways which were plotted based on fold enrichment and gene counts/pathway (dot size), coloured based on -log10(FDR). **C**. Top 10 up- and downregulated proteins per highlighted group. **D**. Heatmap shows the ALSci-specific protein changes with the number of proteins noted. **E**. Gene ontology analysis shows the top enriched molecular terms based on the ALSci-specific proteins. **F**. Schematic figure shows the localization of the ALSci-specific proteins, discovered by the SynGO database. Panel was created using BioRender.com.

Finally, we assessed the role of *C9ORF72*-RE on the ALS synaptic proteome by stratifying BA9 samples based on the patient’s *C9ORF72*-RE status. Analysis of protein changes across both groups (*C9ORF72*-RE-ve; n=12, *C9ORF72*-RE+ve; n=6) revealed almost 600 altered proteins compared to controls (Supplementary Figure 5D, Figure 5A). Interestingly, almost twice as many proteins were altered in the *C9ORF72*-RE+ve group (453) compared to the *C9ORF72*-RE-ve group (257) when compared to controls (Supplementary Figure 5D). KEGG pathway analysis of all 587 altered proteins across groups, revealed enrichment of complement and metabolism pathways again, but also synaptically-important pathways involving different synapse types (glutamatergic and serotonergic), and neurodegenerative disease pathways such as Parkinson’s and Prion disease (Figure 5B). Strikingly, 330 of the 453 altered proteins in the *C9ORF72*-RE+ve group, were specifically altered in that group (Figure 5D). Gene ontology analysis of those 330 proteins revealed significant enrichment of synaptic receptor pathways, with a strong focus on glutamatergic receptor function (Figure 5E). Interestingly, in confirmation of this, subsynaptic analysis of the *C9ORF72*-RE+ve specific proteins revealed that the majority were found postsynaptically and enriched within the postsynaptic density or the actin cytoskeleton (Figure 5F). Strikingly, many key synaptic scaffolding proteins such as SHANKs and Homers were upregulated. In addition, we found that several presynaptic vesicular proteins were altered, which may result in the malfunction of neurotransmitter release. The dramatic postsynaptic changes may be an attempt to compensate for presynaptic dysfunction (Figure 5F).

**Figure 5.**
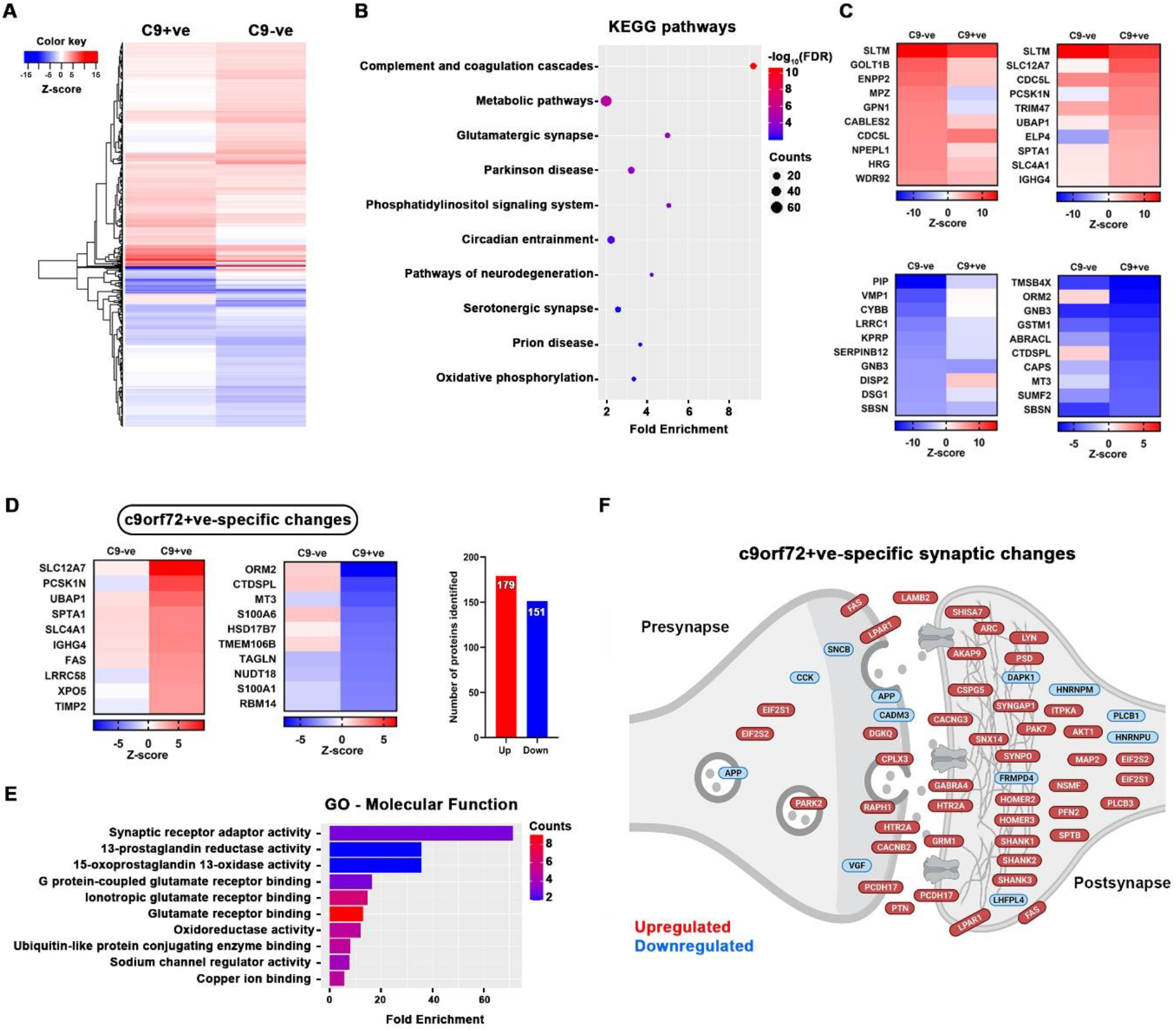
Impact of C9ORF72-RE on ALS synaptic proteome. **A**. Heatmap showing the protein expression differences between C9ORF72-ve and C9ORF72-RE+ve groups. **B**. KEGG pathways revealing the most enriched signalling pathways in both groups. Diagram was plotted based on fold enrichment and gene counts/pathway (dot size), coloured based on - log10(FDR). **C**. Top 10 up- and downregulated proteins per group. **D**. Heatmap shows C9ORF72+ve-specific protein changes with the number of altered proteins highlighted. **E**. Gene ontology analyses show the top enriched molecular terms using C9ORF72+ve-specific protein IDs. **F**. Schematic figure shows the subsynaptic localization of the C9ORF72+ve-specific proteins discovered by the SynGO database. Panel was created using BioRender.com.

To confirm the dramatic postsynaptic change is reflective of changes in the intact brain, we analysed expression and localisation of two proteins at the single synapse level using array tomography. We selected the unchanged presynaptic protein synaptophysin and the upregulated postsynaptic scaffolding protein synaptopodin and compared their expression in 3 randomly selected control, *C9ORF72*-ve and *C9ORF72*+ve cases. Both antibodies produced beautiful synapse-like puncta throughout the BA9 cortical neuropil (Supplementary Figure 6A). Synaptopodin puncta were typically found directly opposed to the presynaptic protein synaptophysin (Supplementary Figure 6B), as expected for a postsynaptic protein [47]. There was no change in synaptophysin density, in agreement with the proteomics data. However, analysis of synaptopodin density revealed an approximate 33% change in *C9ORF72*+ve cases compared to *C9ORF72*-ve cases, which is remarkably similar to the 27% increase observed using proteomics (Supplementary Figure 6C).

## Discussion

In this study we have profiled for the first time, the synaptic proteome in human ALS, revealing regional, genotypic and phenotypic differences. Using an unbiased high-resolution proteomic approach, we have identified hundreds of altered proteins in two key brain areas associated with ALS clinical presentation and assessed the influence of the most common genetic cause of ALS/FTD, *C9ORF72*-RE. Also, we have uniquely validated changes at a single synapse resolution in intact human post-mortem material.

We identified more than 6000 proteins in the human cortical synaptoneurosome fraction. This preparation consists of resealed presynaptic terminals and postsynaptic spines, along with bound glial processes. This provides critical proteomic insight on the full synaptic composition, rather than isolated presynapses or postsynaptic densities, as is common with other synaptic studies. Stringent filtering of our dataset resulted in 5348 analysis-ready proteins which we robustly show are enriched in synapses. However, it is important to consider this to be an enriched and not purified synaptic fraction.

Assessing the changes across the brain in all ALS cases and controls, we identified altered expression of 470 proteins. KEGG pathway analysis highlighted inflammatory processes, synapse subtypes and intriguing ALS-associated pathways such as the spliceosome (Figure 2B,C). Neuroinflammation is a common feature of nearly all neurodegenerative diseases and has been reviewed previously in ALS [48]. Inflammatory proteins at the synapse could appear due to non-autonomous changes in the synaptic milieu or increased synaptic expression of proteins typically considered to be non-neuronal. For example, the complement system is a key feature of the innate immune system [49] but certain components are expressed at synapses, tagging them for removal by microglia [50]. This has recently been shown to be a driver of Alzheimer’s disease pathogenesis [51] and our data revealing increased expression of several complement proteins at the ALS synapse, may suggest a similar process renders ALS synapses vulnerable to aberrant removal. Glutamate excitotoxicity is important in ALS pathogenesis and is driven by excessive activity of excitatory neurons. This could be due to intrinsic changes in neuronal firing properties or reduced inhibitory control by GABAergic cells. It is therefore interesting to note that, “GABAergic synapse” was enriched in our BA4 KEGG analyses and may highlight inhibitory networks as a sight of vulnerability in ALS (Figure 2B). This has been suggested in ALS models and post-mortem human material [52-54]. Alternative splicing of many RNA targets is altered in ALS [55] and key splicing factors and RNA binding proteins are known to function in or near synapses [56, 57]. We find the spliceosome enriched within BA4 ALS synapses, and it would be important to unravel what effect this may have on the splicing and subsequent composition of synaptic proteins, especially given recent data linking TDP-43 mislocalisation to aberrant splicing and decreased expression of the synaptic protein Munc13-1 (UNC13A) [58, 59].

Looking at the individual protein level, there were many changes consistent across cortical regions. For example, SLC12A7 is a K+-Cl− co-transporter (also called KCC4) that maintains intracellular Cl-concentration (critical for balancing neuronal ionic gradients) and plays an important role in regulating cell size by mediating ion transport in response to cell swelling [60]. KCC4 was within the top 10 upregulated proteins in both BA4 and BA9 (Figure 2E), possibly as a shared compensatory mechanism to correct fluctuating neuronal ionic gradients or as a result of cell soma and axonal swelling, a known pathological feature of ALS [61, 62]. Furthermore, GSTM1 (glutathione S-transferase, mu subtype 1) was decreased in both BA4 and BA9 synaptoneurosomes (Figure 2E). GSTM1 is exclusively expressed in astrocytes in mouse brain and is involved in proinflammatory activation of microglia during neuroinflammation [63]. We also observed increased expression of GFAP in BA4 (Figure 2F), highlighting complex bidirectional changes in astrocytic proteins around ALS synapses. This has gained added significance following a recent study highlighting GFAP-containing tripartite synapses as the most vulnerable subtype in ALS spinal cord [64].

### Region-specific alterations

Looking at region-specific changes, we found a significant increased expression of PTP4A3 (also known as PRL-3) in BA4 (Figure 2F). PRL-3 is a phosphatase highly expressed in several cancers, increasing cell proliferation and survival [65]. Very little is known about its physiological role in the brain, however a recent paper [66] highlighted a critical role for the *Drosophila* orthologue (prl-1) in axonal arborisation and synaptogenesis. This may suggest that PRL-3 is significantly upregulated in our data as a compensatory response to retain axonal complexity and synaptic connectivity within a degenerating motor cortex.

Rab34 was significantly and specifically downregulated in BA4 synapses (Figure 2F). Rab34 is a member of the Ras-related proteins in brain (Rab) family, which is involved in intracellular vesicle trafficking and phagosome/lysosome dynamics. Rab34 plays a role in the positioning of lysosomes [67] but is also known to interact with the synaptic vesicle priming protein Munc13-2 [68, 69]. Munc13-2 is important for vesicle priming and is often co-expressed in the same synapses as Munc13-1 [70]. Mutations in the Munc13-1 gene (UNC13A) are causative for ALS and two recent papers highlight TDP-43-dependent alterations in UNC13A and decreased protein product [58, 59]. Therefore, decreased expression of Rab34 at BA4 synapses may render these vulnerable due to inefficient vesicle priming and disrupted synaptic transmission. We discovered Munc13-1 at the synapse but did not see a change in expression level, likely due to the mixed population of synapses collected from neurons with/without TDP-43 mislocalisation.

In the frontal cortex, TRIM47 was specifically upregulated more than 2-fold compared to control synapses (Figure 2H). TRIM47 is an E3 ubiquitin-protein ligase that mediates the degradation of CYLD lysine 63 deubiquitinase (CYLD) [71], which has very recently been highlighted as a rare causative gene for FTD [72, 73] and possibly ALS [74]. Interestingly, CYLD is known to interact with proteins already implicated in ALS/FTD, such as TBK1, OPTN and SQSTM1 [72]. Few cases have been described and there is debate around what effect the disease-associated mutations have on CYLD activity [74, 75], however it is interesting to note that the most upregulated protein in the ALS frontal cortex is known to regulate levels of a protein directly implicated in FTD. Supporting the link between TRIM47 and cognitive change, TRIM47 was increased more than 2-fold in the ALSci samples. TRIM47 has also been recently shown to accelerate brain injury after cerebral ischaemia/reperfusion, with TRIM47 knockdown being both neuroprotective and anti-inflammatory [76]. Therefore, increased TRIM47 expression may indicate inflammatory processes at play within the synaptic milieu.

PCP4 (also known as PEP-19) is a calcium/calmodulin binding protein which regulates calmodulin function by modulating its interaction with calcium [77]. PEP-19 is expressed in the cortex of post-mortem human brain and its levels are significantly decreased in Alzheimer’s, Huntington’s and Parkinson’s disease [78]. PEP-19 knockout in mice produces significant alterations in synaptic plasticity and behaviour, likely due to its loss of control over calmodulin activity, a key process in synaptic plasticity [79]. Here, we discovered a specific decrease of PEP-19 in synapses of the ALS frontal cortex (Figure 2H), introducing PEP-19 as a potential new player in ALS synaptic dysfunction.

### Synaptic alterations in ALSci

ALSci patients have a worse prognosis and present with cognitive and behavioural changes, reminiscent of FTD [46]. We recently discovered that ALSci cases have synapse loss in the frontal cortex [12] and in this study we stratified our samples by cognitive status to try and uncover the molecular changes that may influence synaptic integrity. We identified 143 proteins specifically altered in the ALSci BA9 samples and gene ontology analysis revealed enrichment of toll-like receptor 4 (TLR4) pathways (Figure 3E), critical components of the innate immune system. Recent work has shown TLR4-dependent synapse loss after traumatic brain injury, which could be reduced by TLR4 blockade [80]. Furthermore, TLR4 knockout in hSOD1G93A mice leads to improved grip strength and extended survival [81]. TLR4 induction also leads to increased synapse loss around spinal motor neurons in peripheral nerve crush models [82]. Together, this suggests our observation of TLR4 pathway enrichment is likely to be detrimental to synaptic function and structure.

RNF146 (also known as Iduna) is an E3 ubiquitin ligase that uses poly (ADP ribose) (PAR) to recognise its substrates [83], tagging them for degradation, and here is specifically upregulated in ALSci synapses (Figure 3D). PAR polymerases (PARPs) add PAR motifs to target proteins, with PARP1 being the most prolific [84]. PARP1 initiates a form of cell death known as parthanatos [85] and recent studies describe increased PARP1 activity in ALS spinal motor neurons and the beneficial effects of a small molecule PARP inhibitor [86, 87]. Interestingly, Iduna is an NMDA-receptor induced survival molecule that interferes with parthanatos [88]. Iduna is normally expressed at very low levels in neurons, but following NMDA receptor activation or neuronal stress, Iduna levels significantly increase and have a neuroprotective effect [88]. Therefore, increased RNF146 expression may represent a pro-survival signal retained within stressed synapses, or disease associated RNF146 increase may drive ubiquitination and subsequent degradation of important synaptic proteins.

FRMPD4 was the most significantly decreased protein in ALSci BA9 (Figure 3D). FRMPD4 is a PSD-95 binding protein that regulates dendritic spine morphogenesis and interacts with Homer and mGluR1/5 to regulate downstream signalling [89, 90]. Furthermore, mutations in FRMPD4, resulting in significant protein loss, have been linked to a severe form of intellectual disability [91]. Interestingly, a recent transcriptomic study in the frontal cortex of sporadic FTD cases, identified a decrease in FRMPD4 expression [92]. Together, this advances the cognitive and pathological overlap between ALSci and FTD to the synaptic level, and highlights FRMPD4 as a key synaptic protein affected in both diseases.

Given the worse prognosis of ALSci patients, we hypothesised these patients may have a more aggressive form of disease, resulting in a distinct molecular change in their BA4 synapses. We discovered 230 specifically altered proteins in the ALSci BA4 samples, and gene ontology analysis highlighted strong enrichment of RAGE receptor pathways (Figure 4E). In fact, numerous RAGE receptor ligands were upregulated in BA4 synapses, including several S100 calcium-binding proteins. RAGE receptor activation leads to the strong induction of pro-inflammatory signals and increased expression has been shown at presymptomatic stages of SOD1 and TDP43 mouse models of ALS [93]. Furthermore, RAGE knockout in hSOD1G93A mice leads to reduced inflammation and extended lifespan [93]. In a recent human dataset a negative association between high RAGE expression and earlier age at death was observed [94], supporting our idea that ALSci patients have a more severe form of disease, and this may be driven by RAGE-dependent mechanisms.

Interestingly, we observed a significant decrease in synaptic expression of HTR1A, (the 5HT1A serotonin receptor) in BA4 of ALSci samples (Figure 4D). Previous human PET imaging revealed a decrease in 5HT1A receptor binding in the cortex (motor and extra-motor regions) of ALS patients versus controls [95]. ALS patients were not tested for cognitive impairment, so data cannot be stratified to assess any association between ALSci and 5HTR1A reduction. Here we identified a loss of 5HT1A receptor in the motor cortex of ALSci patients, but surprisingly found an increased expression in BA9. This may reflect synaptic changes at different disease stages, as the disease spreads forward from motor to non-motor regions.

### The impact of C9ORF72-RE on synaptic proteome

The physiological role of the C9ORF72 protein is not fully understood, but it is believed to regulate vesicle trafficking and autophagy by interacting with Rab proteins [96]. Furthermore, recent studies have revealed C9ORF72 to be presynaptic and to interact with Rabs involved in synaptic vesicle trafficking [43, 97, 98]. This has been reinforced in iPSC-derived neurons from *C9ORF72*-RE patients, showing presynaptic dysfunction and altered neuronal network activity [99, 100]. Given the growing literature on the synaptic localisation and function of C9ORF72, we stratified our BA9 samples by the patient’s *C9ORF72*-RE status. There were 587 altered proteins, regardless of *C9ORF72*-RE status and KEGG pathway analysis highlighted enrichment of inflammatory pathways, but also synaptic subtypes (glutamatergic and serotonergic) and several disease-associated pathways such as Parkinson’s disease and Prion disease (Figure 5B). 453 proteins were altered in the *C9ORF72*-RE+ve group, of which a remarkable 330 were specifically altered (Figure 5D). Interestingly, gene ontology analysis highlighted enrichment of synaptic receptor pathways, specifically post-synaptic glutamatergic pathways (Figure 5E), reinforcing the influence of glutamate-induced hyperexcitability in *C9ORF72-*associated ALS [101]. Metabotropic glutamate receptor 1 and important scaffolding proteins (SynGAP1, SHANKs and Homers) were all upregulated (Figure 5F). This dramatic postsynaptic reorganisation may be a response to altered presynaptic function. We discovered several proteins involved in vesicular trafficking and homeostasis to be altered at the presynapse. It fits that decreased expression of C9ORF72 protein as a result of *C9ORF72*-RE, may impact presynaptic function which results in a compensatory reorganisation of the postsynapse to try and maintain synaptic balance. We have also shown for the first time at the single-synapse level in intact human tissue, that postsynaptic scaffolding proteins such as synaptopodin are upregulated in *C9ORF72*-RE+ve cases (Supplementary Figure 6). Furthermore, increased expression of synaptotagmin-13 is of note, due to recent work suggesting this presynaptic vesicular protein is preferentially expressed in resilient neurons in ALS and Syt13 gene therapy increased survival of SOD1G93A ALS mouse models [102]. These results highlight the complexity of compensatory pre and postsynaptic protein alterations to try and maintain synaptic integrity.

Increasing more than 2-fold was PCSK1N, also known as pro-SAAS (Figure 5D). This small secretory chaperone is expressed throughout the brain [103] and has been linked to several neurodegenerative diseases, including FTD [104-107]. Pro-SAAS can prevent fibrillisation of disease associated proteins such as amyloid [108] and a-synuclein [109], suggesting it may serve important neuroprotective roles. Importantly, several ALS-associated proteins undergo fibrillisation and subsequent aggregation, such as TDP-43 [110] and the dipeptide repeats (DPRs) generated from *C9ORF72*-RE [111]. Therefore, this may suggest that pro-SAAS upregulation at the *C9ORF72*-RE+ve synapses is an attempt to combat fibrillisation and aggregation of disease-associated proteins.

Strikingly, TMEM106B was specifically decreased in the *C9ORF72*-RE+ve samples (Figure 5D). TMEM106B is involved in lysosomal homeostasis and overexpression leads to abnormalities in lysosmal acidification, size and number [112]. *TMEM106B* is a genetic risk factor for FTLD-TDP, especially in patients with granulin mutations [113], yet intriguingly the same alleles appear protective for *C9ORF72*-RE carriers, resulting in later age of onset and death [114]. Furthermore, TMEM106B knockout in a granulin model of ALS, leads to reversal of lysosomal alterations and prevention of neurodegeneration [115]. Therefore, specific reduction of TMEM106B in *C9ORF72*-RE+ve samples may be a compensatory attempt to regulate lysosomal function, or a concomitant loss of lysosomal proteins due to *C9ORF72*-RE-dependent lysosomal dysfunction. Alternatively, lower synaptic expression of TMEM106B may be due to mislocalisation and aggregation in other neuronal compartments, as recently shown in FTLD-TDP [116]. Finally, TMEM106B knockdown leads to altered lysosomal trafficking and reduced dendritic arborisation [117], so a loss of TMEM106B at the synapse may drive vulnerability.

### Summary

In summary, we have mapped the first unbiased proteome of the human ALS synapse, uncovering novel players in synaptic dysfunction and highlighting ALS-associated proteins in this vulnerable subcellular compartment. Future work in model systems will uncover the mechanisms leading to these protein changes, adding critical insight into ALS pathogenesis, and highlighting novel avenues of understanding and therapeutic development.

## Data Availability

All data produced in the present study are available upon reasonable request to the authors and will be deposited on publicly available repositories once published.

## Acknowledgements

We gratefully acknowledge the generous donors who have made this work possible. We wish to acknowledge the CARE-MND Register, hosted by the Euan Macdonald Centre (EMC) for MND Research and funded by MND Scotland. We are extremely thankful for the hard work of the Scottish MND Clinical Specialist Team for obtaining consent for tissue donation, led by Judith Newton. **CMH** acknowledges funding support from MND Scotland, TENOVUS Scotland and Alzheimer’s Research UK (ARUK-EG2019B-003). **ZIL** holds an MND Association Lady Edith-Wolfson Junior Fellowship. **ASA** is funded by a Euan Macdonald Centre for Motor Neuron Disease Research PhD studentship. **TSJ** receives funding from the European Research Council (ERC) under the European Union’s Horizon 2020 research and innovation programme (grant agreement no. 681181) and the UK Dementia Research Institute which receives its funding from DRI Ltd, funded by the UK Medical Research Council, Alzheimer’s Society, and Alzheimer’s Research UK. **RAK** was funded by the Euan Macdonald Centre for Motor Neuron Disease Research PhD studentship, and the University of Edinburgh’s Principal’s Career Development and Global Research scholarships. **TMW** and **SLE** are supported by funding from the Biotechnology and Biological Sciences Research Council (BBSRC) Institute Strategic Grant (BBS/E/D/10002071). We thank Dominic Kurian from the Roslin Institute Proteomic and Metabolomic Facility for advice.

---

**Supplementary Figure 1.**
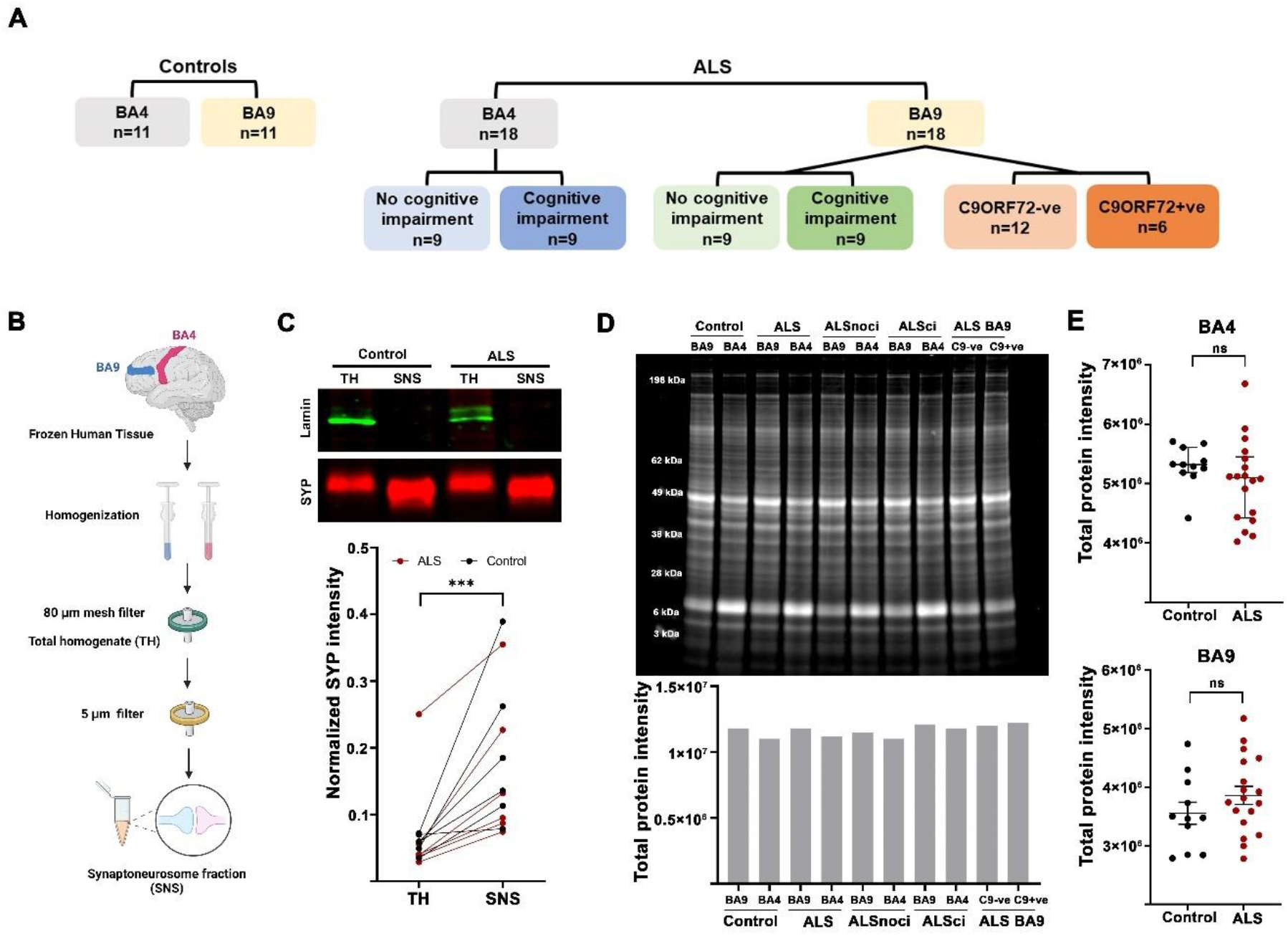
Graphical summary of experimental plan and synaptic preparations. **A**. Experimental strategy, highlighting stratification and grouping of all samples. Each coloured shape represents an experimental group, and the n-number highlights the number of cases within that group. **B**. Graphical representation of the synapse enrichment protocol. **C**. Western blots from control and ALS samples confirm exclusion of nuclear protein (lamin) and enrichment of synaptic protein (synaptophysin) in synaptoneurosome (SNS) preps. Paired t-test, ***p<0.0001. **D**. Total protein Coomassie stain of a gel containing 5ug of each group sample shows similar protein intensity and banding patterns. **E**. Quantification reveals no difference in total protein levels between individual control and ALS samples in both brain regions. BA4: Mann-Whitney test, p= 0.1015, BA9: Unpaired t-test p=0.2293. Graphs show individual cases per group (dots) and median ± IQR or mean ± SEM.

**Supplementary Figure 2:**
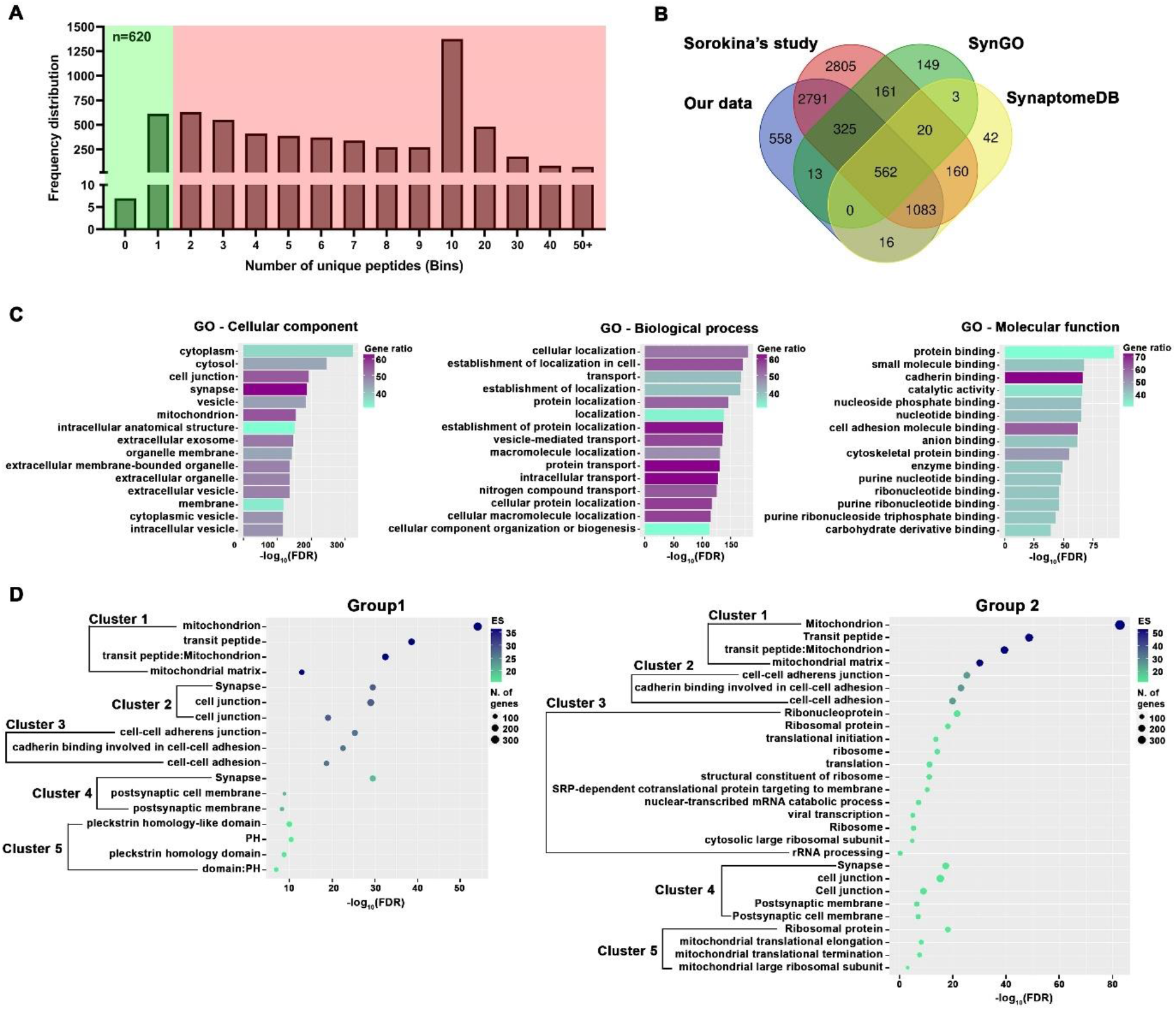
Further enrichment analysis of our synaptic samples. **A**. All protein IDs derived from fewer than two unique peptides (n=620) were excluded from further analysis. **B**. Filtered, final dataset was aligned with 3 different synaptic databases, and showed ∼90% overlap, thereby validating the composition of our synaptic fractions. **C**. Enrichment analysis was performed using g:Profiler. Gene Ontology (GO) clusters were plotted using the top 15 biological terms, ranked by -log10(FDR) and coloured by gene ratio (gene number/term size). **D**. Functional annotation clustering was performed using DAVID Functional Annotation Bioinformatics Microarray Analysis software. Our large dataset was halved using a random group generator (DAVID only accepts inputs of 3000 IDs) and each group was uploaded into DAVID and analysed. Both plots represent the top 5 clusters with different biological terms, plotted by -log10(FDR) and gene count (dot size), coloured based on the DAVID enrichment score per cluster.

**Supplementary Figure 3:**
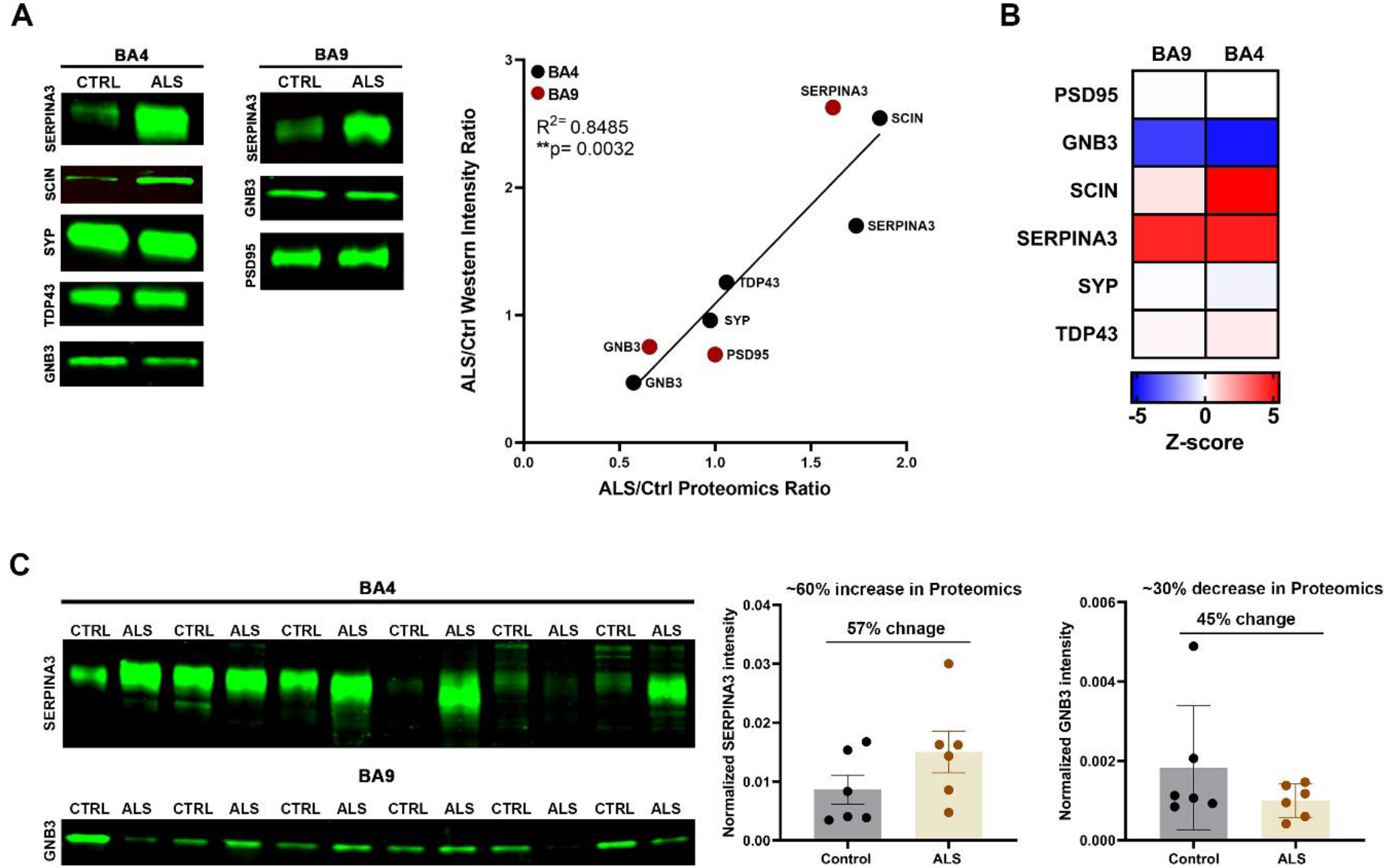
Comparison of proteomics data with western blot analyses. **A**. Representative western blot images using BA4 and BA9 pooled samples shows significant correlation with proteomics results. Control normalized proteomics intensity values (corrected by 1/median) were plotted against control normalized western blot intensity values. Pearson’s correlation, R^2^=0.8485, **p=0.0032. **B**. Heatmap representation of proteins in **(A). C**. Western blot images show SERPINA3 and GNB3 expression in individual cases from different brain areas. Although the quantification shows no significant difference between Ctrl and ALS groups, there is a coherence with the proteomics data. SERPINA3: Unpaired t-test, p=0.1681; GNB3: Mann-Whitney test, p=0.5887. Graphs show mean ± SEM or median ± IQR.

**Supplementary Figure 4.**
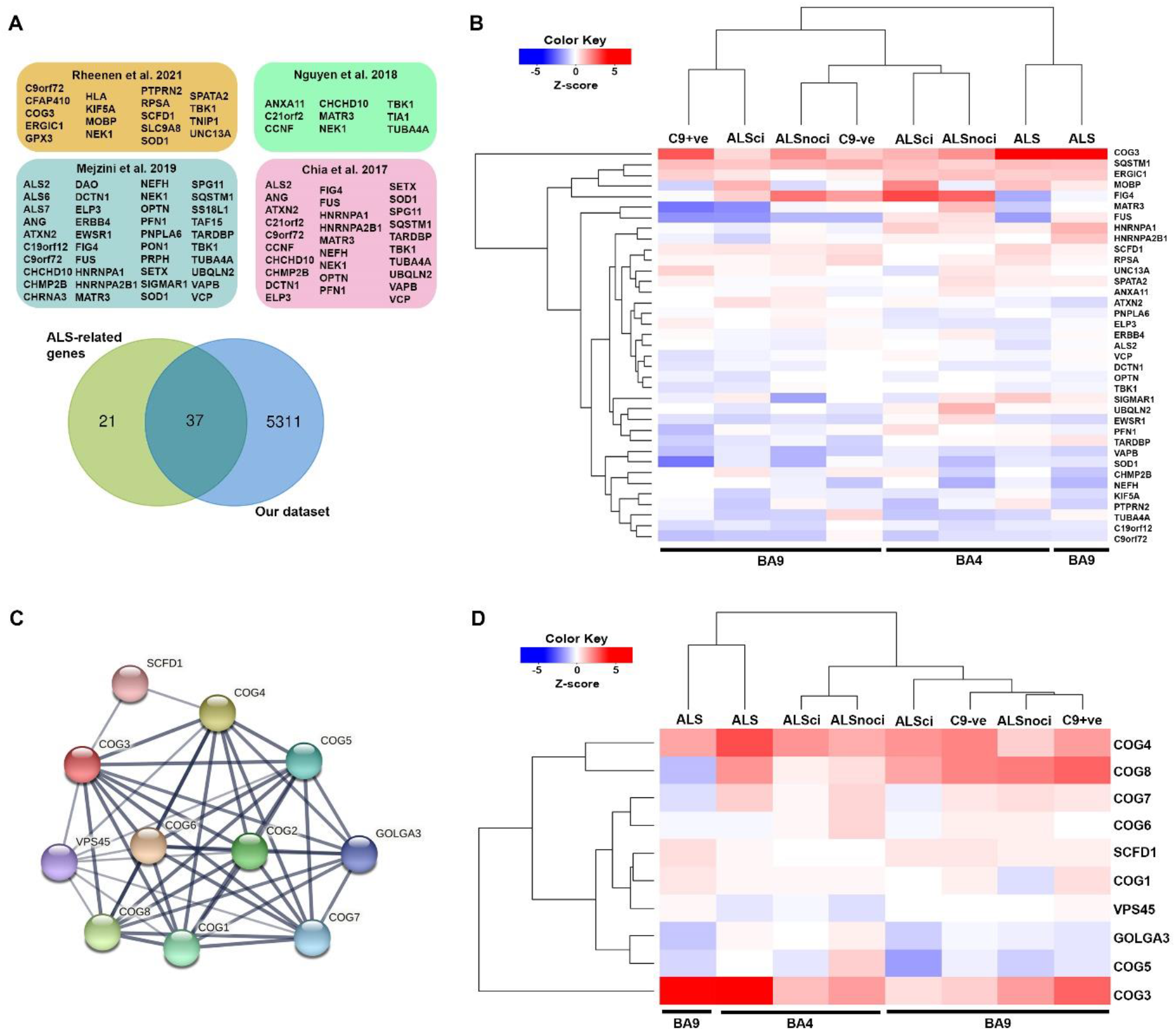
Expression profile of ALS-related genes. **A**. 58 ALS-related genes were collated from the 4 highlighted references. **B**. Heatmap showing the differences in expression of the protein product from the 37 ALS-related genes found in our dataset. **C**. Interaction map of the most upregulated gene, COG3. Diagram was made using the STRING database, edges indicate that the proteins are part of a physical complex, meanwhile the line thickness indicates the strength of supporting data. **D**. Heatmap showing the protein expression profile of the COG3-based network highlighted in **C**. Heatmaps was made using hierarchical clustering with Euclidean distance.

**Supplementary Figure 5:**
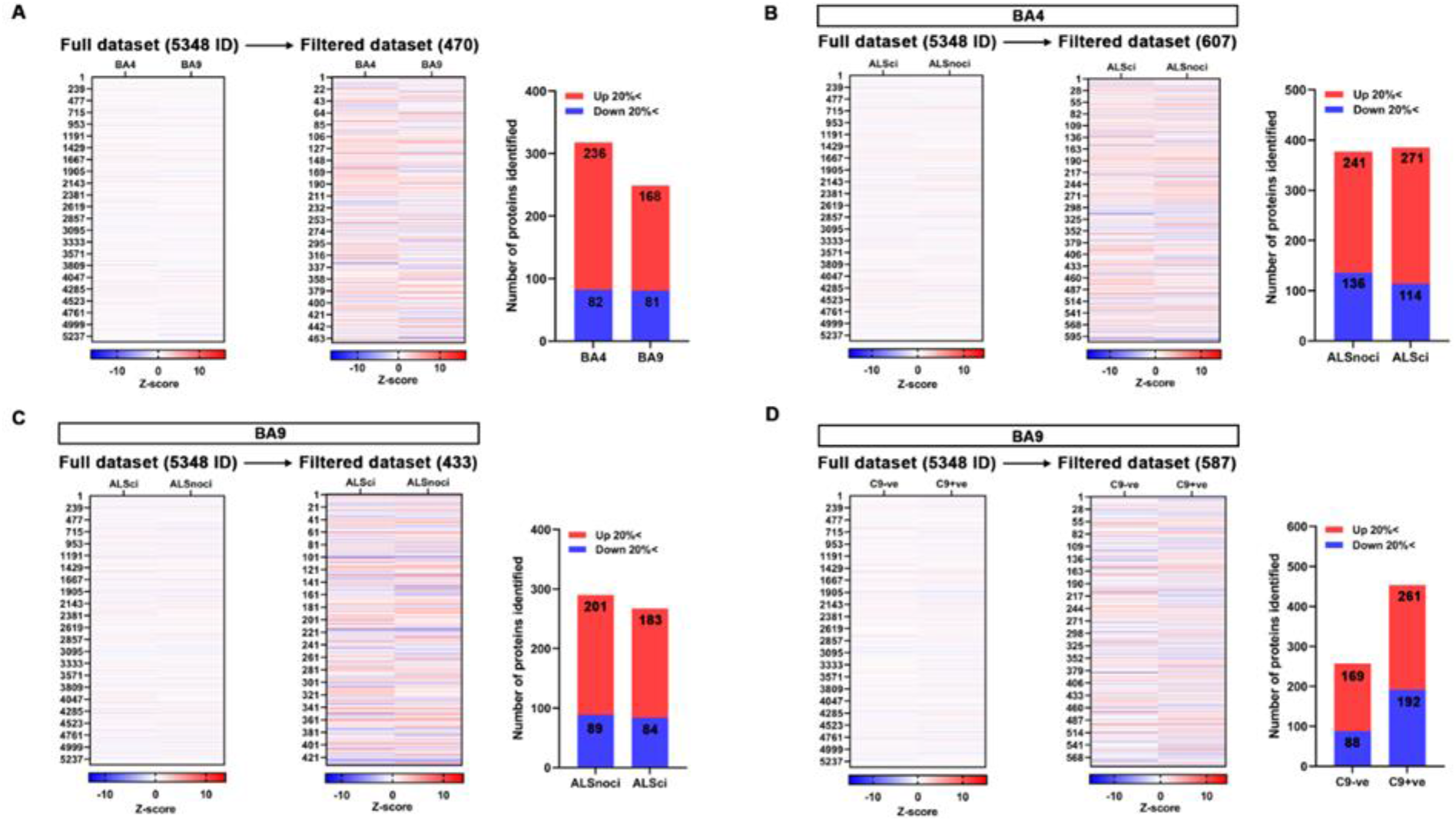
Data filtration to identify proteins for further analysis. **A-D**. Heatmaps show the filtration process of the data in each experimental comparison. The first heatmap represents the expression change of all 5348 proteins versus control samples, and the second represents the proteins that have altered expression (± 20%) versus control. Bar graphs represent the number of up- and downregulated proteins in each group versus control. Data was stratified by brain region (**A**), cognitive performance (**B** and **C**) and genetic status (**D**).

**Supplementary Figure 6.**
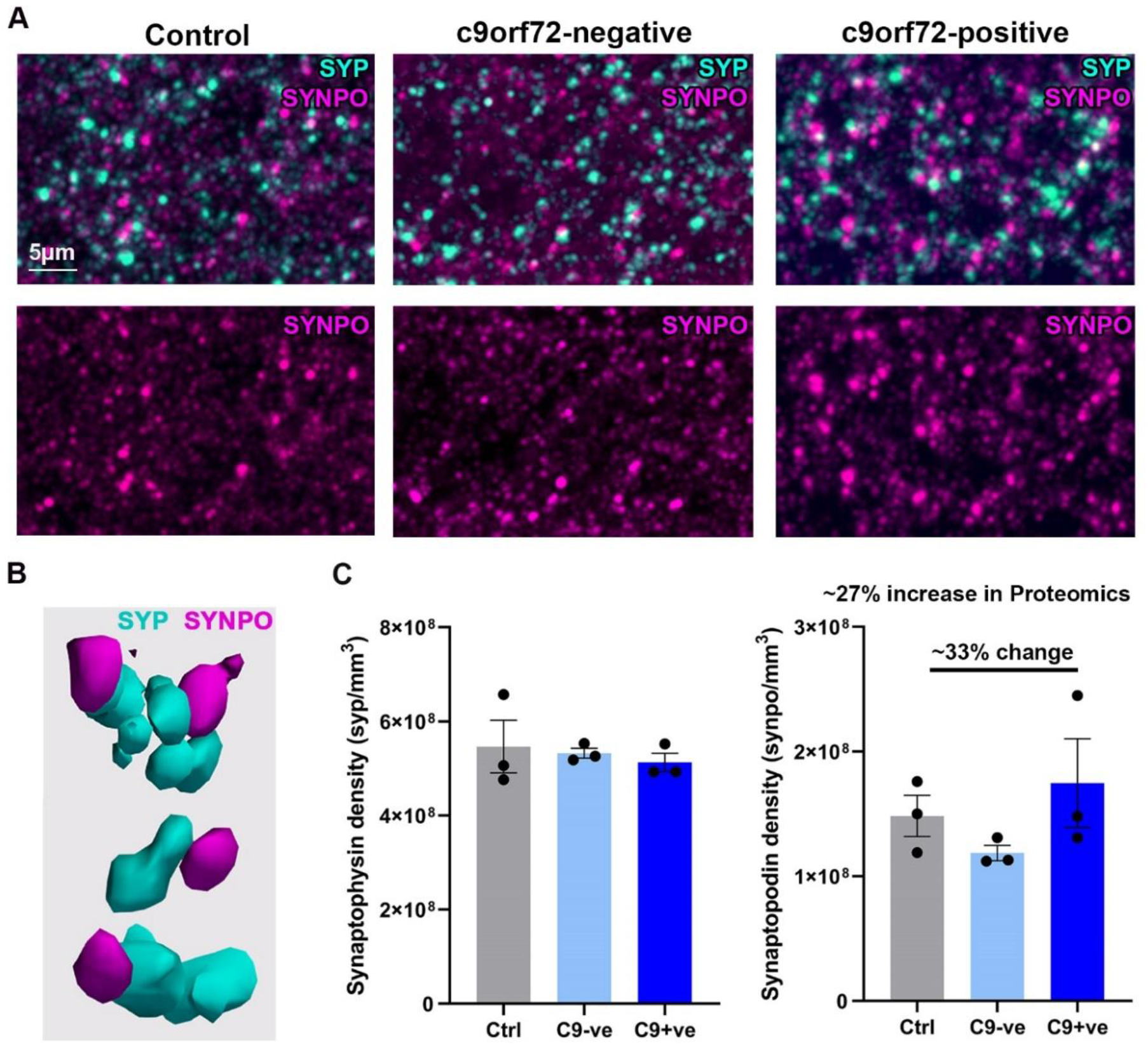
Analysing protein change at single synapse resolution. **A**. Example micrographs of synaptophysin (SYP) and synaptopodin (SYNPO) staining of post-mortem human tissue sections using array tomography. Single channel images suggest increased density of SYNPO puncta in c9or72-RE+ve tissue. **B**. Synapses consisting of presynaptic SYP and postsynaptic SYNPO were consistent throughout the neuropil. This example was obtained following segmentation and 3D rendering of array tomography images which were used for quantification, using the free imaging tool Paraview (paraview.org). **C**. Graphs show the density of SYP or SYNPO in each group using 3 cases per group. SYNPO shows a similar change in C9ORF72-RE+ve samples that ratiometric proteomics data showed. Groups were compared with one-way ANOVA using Tukey’s multiple comparison test, no significant difference were found. Graphs show mean±SEM.

**Supplementary Table 1.** Primary and secondary antibodies used in the study. Details of antibody, manufacturer and working dilution, plus secondary antibodies used for western blots and array tomography.

| Primary antibody | Manufacturer | Species | Dilution | Secondary antibody |
| --- | --- | --- | --- | --- |
| **Synaptophysin** | Abcam<br>(ab8049) | Mouse | 1:1000 WB<br>1:50 AT | WB: Donkey anti-Mouse IgG IRDye® 800CW, Li-Cor (923-32212)<br>AT: Donkey anti-mouse Alexa fluor 488 Abcam (ab150105) |
| **Lamin** | Proteintech<br>(10298-1-AP) | Rabbit | 1:1000 | Donkey anti-Rabbit IgG IRDye® 800CW, Li-Cor (926-32213) |
| **SERPINA3** | Proteintech<br>(12192-1-AP) | Rabbit | 1:1000 | Donkey anti-Rabbit IgG IRDye® 800CW, Li-Cor (926-32213) |
| **SCIN** | Proteintech<br>(11579-1-AP) | Rabbit | 1:1000 | Donkey anti-Rabbit IgG IRDye® 800CW, Li-Cor (926-32213) |
| **TDP-43** | Abcam<br>(ab133547) | Rabbit | 1:1000 | Donkey anti-Rabbit IgG IRDye® 800CW, Li-Cor (926-32213) |
| **GNB3** | Proteintech<br>(10081-1-AP) | Rabbit | 1:1000 | Donkey anti-Rabbit IgG IRDye® 800CW, Li-Cor (926-32213) |
| **PSD95** | Cell Signalling<br>(3450S) | Rabbit | 1:1000 | Donkey anti-Rabbit IgG IRDye® 800CW,<br>Li-Cor (926-32213) |
| **SYNPO** | Proteintech<br>(21064-1-AP) | Rabbit | 1:50 | Donkey anti-rabbit Alexa fluor 594 –<br>Abcam (ab150076) |

